# Indoor transmission of SARS-CoV-2

**DOI:** 10.1101/2020.04.04.20053058

**Authors:** Hua Qian, Te Miao, Li Liu, Xiaohong Zheng, Danting Luo, Yuguo Li

**Affiliations:** School of Energy and Environment, Southeast University, Nanjing, China; Department of Mechanical Engineering, The University of Hong Kong, Pokfulam, Hong Kong, China; School of Architecture, Tsinghua University, Beijing, China; School of Public Health, The University of Hong Kong, Pokfulam, Hong Kong, China

**Keywords:** COVID-19, indoor environments, indoor hygiene

## Abstract

**Background:** By early April 2020, the COVID-19 pandemic had infected nearly one million people and had spread to nearly all countries worldwide. It is essential to understand where and how SARS-CoV-2 is transmitted.

**Methods:** Case reports were extracted from the local Municipal Health Commissions of 320 prefectural cities (municipalities) in China, not including Hubei province, between 4 January and 11 February 2020. We identified all outbreaks involving three or more cases and reviewed the major characteristics of the enclosed spaces in which the outbreaks were reported and associated indoor environmental issues.

**Results:** Three hundred and eighteen outbreaks with three or more cases were identified, involving 1245 confirmed cases in 120 prefectural cities. We divided the venues in which the outbreaks occurred into six categories: homes, transport, food, entertainment, shopping, and miscellaneous. Among the identified outbreaks, 53·8% involved three cases, 26·4% involved four cases, and only 1·6% involved ten or more cases. Home outbreaks were the dominant category (254 of 318 outbreaks; 79·9%), followed by transport (108; 34·0%; note that many outbreaks involved more than one venue category). Most home outbreaks involved three to five cases. We identified only a single outbreak in an outdoor environment, which involved two cases.

**Conclusions:** All identified outbreaks of three or more cases occurred in an indoor environment, which confirms that sharing indoor space is a major SARS-CoV-2 infection risk.

**Funding:** The work was supported by the Research Grants Council of Hong (no 17202719, C7025-16G), and National Natural Science Foundation of China (no 41977370).

## INTRODUCTION

In less than 4 months, SARS-CoV-2 has rapidly spread to nearly all countries worldwide and by 3 April 2020 it had infected more than a million people, and killed nearly 50,000 people.^1^ Understanding where and how SARS-CoV-2 is transmitted from an infected person to a susceptible person is essential for effective intervention.

The once-in-a-century COVID-2019 pandemic occurred right in the age of artificial intelligence and big data. Many clusters/outbreaks were identified via contact tracing by local health authorities in China and elsewhere using both traditional and new technologies. The identification of these clusters allowed the health authorities to quarantine close contacts for effective intervention and provided an opportunity to study the characteristics of where and how these clusters occurred. The first COVID-2019 patient was identified in Wuhan in December 2019, and the largest number of confirmed Chinese cases occurred in Hubei province, of which Wuhan is the provincial capital.^2^ Since 20 January 2020, the local health authorities of cities outside Hubei have reported online the details of most identified cases of infections.

In this study, we identified the outbreaks from these case reports from the local Municipal Health Commissions of 320 prefectural cities (municipalities) in China, not including Hubei province, between 4 January and 11 February 2020 and reviewed the major characteristics of the enclosed areas in which these outbreaks were determined to have occurred and associated indoor environment issues.

## Methods

We collected descriptions of each confirmed case from the local Municipal Health Commission website of 320 prefectural cities in mainland China, not including Hubei province. Each local Municipal Health Commission announced a description of the confirmed cases each day. The case descriptions generally included age, sex, venue of infection, symptoms, date of symptom onset, hospitalisation, and confirmation and history of exposure. Many described cases also included the individual trajectory and relationship with other confirmed cases, and quite often clusters had already been identified. We consulted the websites nationwide except for those of cities in Hubei province and collected all available data up to 11 February 2020. Data from a few major cities – Beijing, Shanghai, and Guangzhou – were not included in our analysis due to insufficient case descriptions. Case descriptions from Hong Kong, Macao, and Taiwan were collected from their health authorities. We input the data into a database in a unified format and conducted cross-validation to ensure data reliability.

A total of 7324 cases with the minimum required descriptions (i.e., the information listed above) were found; these accounted for 66.7% of the 10,980 confirmed non-Hubei cases in China by 11 February 2020.^3^ In this study, we defined a *cluster* as an aggregation of *three* or more cases that appears to be linked to the same infection venue (e.g., an apartment, an office, a school or a train) during a sufficiently close period. We defined an *outbreak* as a cluster in which a common index patient is suspected, and we excluded tertiary and higher-generation infections in counting the number of cases involved. We also excluded outbreaks that involved only two cases to exclude possible spouse-to-spouse transmission and to reduce the workload due to the large number of clusters or outbreaks with two cases. We also identified the index patient(s) of the identified outbreaks and their date of symptom onset.

We divided the identified outbreaks into categories for further analysis. First, six categories of infection venues were considered: homes (apartments and villas), transport (train, private car, high-speed rail, bus, passenger plane, taxi, cruise ship, etc.), restaurants and other food venues, entertainment venues (gyms, mahjong, cards, tea houses, and barbershops) and shopping venues (shopping malls and supermarkets), with an additional miscellaneous venue (hospitals, hotel rooms, unspecified community, thermal power plants, etc.). Second, four categories of infected individuals were considered based on their relationship: family members, family relatives, socially connected and socially non-connected.

### Role of the funding source

The funding bodies had no such involvement in study design; in the collection, analysis, and interpretation of data; in the writing of the report; and in the decision to submit the paper for publication.

The corresponding authors also confirms that they had full access to all the data in the study and had final responsibility for the decision to submit for publication.

## Results

We identified 318 outbreaks involving 1,245 infected individuals in 120 cities. The top three cities (Table S2) were Shenzhen, Guangdong (24 outbreaks, 7·5%; 84 cases, 6·7%), Chongqing (16 outbreaks, 5·0%; 61 cases, 4·9%) and Bozhou, Anhui (nine outbreaks, 2·8%; 35, 2·8%). The average number (±SD) of cases per outbreak was 3·92±1·65. Among the 318 identified outbreaks, more than half (171; 53·8%) involved three cases, more than a quarter (84; 26·4%) involved four cases, and only five (1·6%) outbreaks involved ten or more cases. Table S1 briefly describes four outbreaks, including the largest outbreak in a shopping mall in Tianjin (21 cases).

Among the 318 outbreaks, 129 involved only family members, 133 involved family relatives, 29 involved socially connected individuals, 24 involved socially non-connected, and only three involved multiple relationships. In addition to family members, family relatives and socially connected individuals constituted a large proportion of the infected cases (Figure 1A).

**Figure 1.**
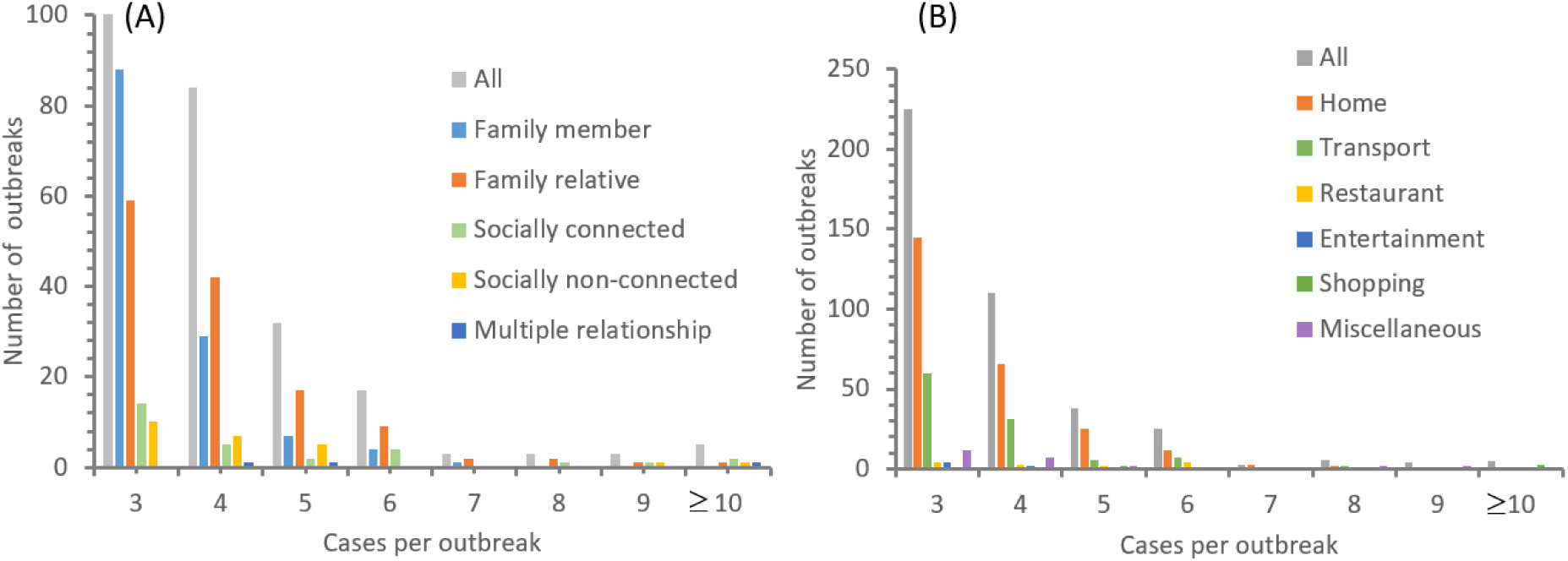
Distribution of all identified 318 outbreaks. (A) involving confirmed cases of different relationships and (B) for each category of the 416 venues.

Eighty-three of the 318 identified outbreaks had multiple possible venues, which means either that the exact venue of infection cannot be identified or that more than one venue was involved in the infection. If we double- or triple-count these venues, we have a total of 416 infection venues for 318 outbreaks (Figure 1B). Among the 318 outbreaks, 254 (79·9%) occurred in a home (one in a villa; all others in apartments), 108 (34·0%) in transport, 14 at a restaurant or other food venue, seven at an entertainment venue, and seven at a shopping venue (shopping mall and supermarket), with an additional 26 at a miscellaneous venue (e.g., hospital, hotel room, unspecified community, and thermal power plant).

Most of the 254 home outbreaks included three to five cases (145 with three cases, 66 with four cases, and 25 with five cases). The average number of cases was 3·7 for the home outbreaks, 3·8 for transport, 4·9 for food venues, 3·6 for entertainment venues, 8·7 for shopping venues, and 4·4 for miscellaneous venues. The proportion of large outbreaks was high for shops and food venues, possibly because more susceptible individuals were present in these venues than in homes. Shopping and entertainment venues were each associated with only seven outbreaks. This seems to suggest the difficulty of implementing preventive measures in places with large numbers of susceptible individuals.

Between 29 December 2019 and 31 January 2020, we also identified 231 outbreaks with known start and end dates of the suspected infectious period (Figure 2A). The identified outbreaks peaked between 23 and 28 January (Figure 2A), which coincides with the celebration period of the Chinese New Year (CNY). CNY 2020 lasted from New Year’s Eve on 24 January to the Lantern Festival (i.e., the 15^th^ of Lunar January) on 8 February. The official holiday in mainland China was from 24 to 30 January 2020. The peak date for the number of transport outbreaks was 1 to 2 days earlier than that for the home outbreaks as people travelled home for CNY.

**Figure 2.**
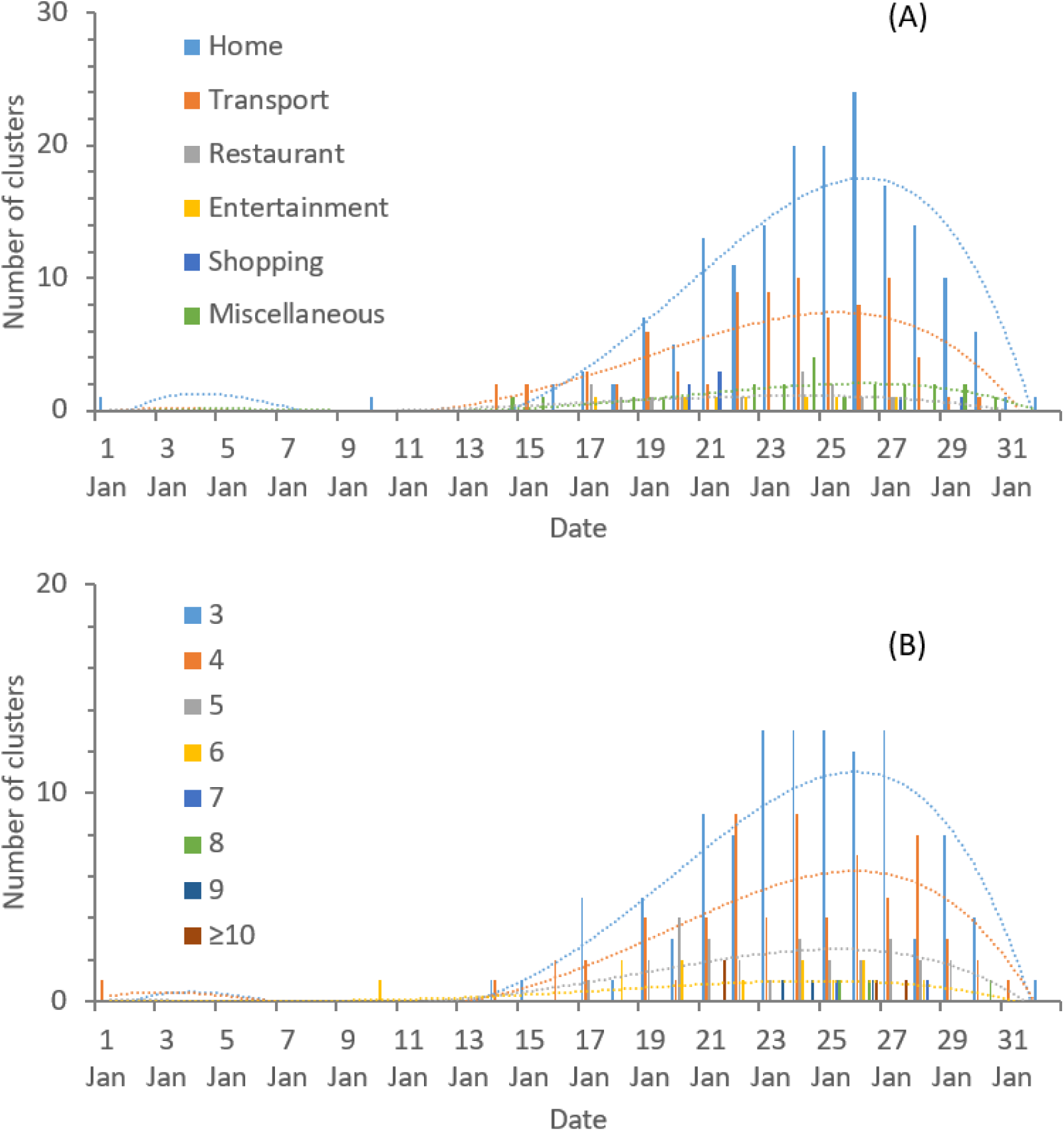
Occurrence of outbreaks on median dates of infectious period for 231 actual outbreaks. (A) in different categories for 300 venues and (B) with different numbers of cases in each outbreak on median dates of infectious period.

Because home outbreaks dominated, the changes in the temporal profile of the number of cases (Figure 2A) closely follows that of the home outbreaks (Figure 2B). However, for outbreaks with more than six cases, no particular pattern was identified over time, which suggests a sporadic nature.

Among the 231 outbreaks with a known suspected infectious period, 126 included a known date of symptom onset for the index patient (Figure 3). We further divided those 126 outbreaks into two subgroups according to the index patient’s symptom-onset date: on or before 28 January (96 outbreaks) and after 28 January (30 outbreaks). The average time from symptom onset to the ending infectious date was 3·76±4·42 days for those on and before 28 January and 0·87±2·80 days for those afterward.

**Figure 3.**
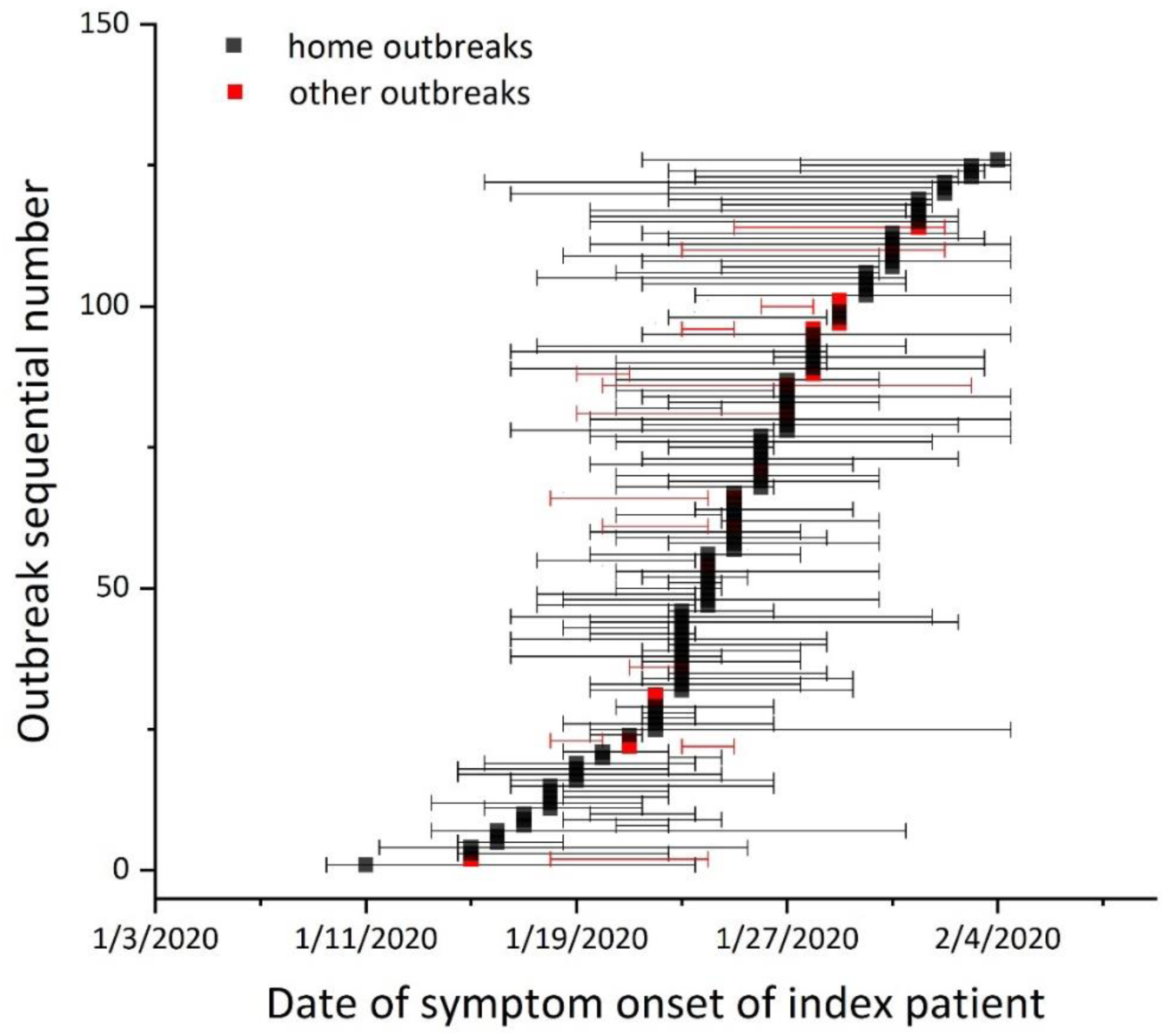
One hundred and twenty-six outbreaks with both possible starting and end infectious dates and date of symptom onset of index patient. Non-home outbreaks are shown in red.

## Discussion

The first salient feature of the 318 identified outbreaks that involved three or more cases is that they all occurred in indoor environments. Although this finding was expected, its significance has not been well recognised by the community and by policy makers. Indoors is where our lives and work are in modern civilisation. The transmission of respiratory infections such as SARS-CoV-2 from the infected to the susceptible is an indoor phenomenon.

The emergence of homes as the most common COVID-19 outbreak venue in China is not surprising. During the COVID-19 epidemic in mainland China, homes became temporary quarantine places. Our estimated home dominance of 79·9% is close to the official estimate of 83% of the so-called household clusters among the nearly 1000 clusters (not outbreaks) defined by the China National Health Commission.^4^ After Wuhan announced its city lockdown on 23 January, the warning message spread throughout the country. People in provinces outside Hubei also began to stay at home. Most Chinese families have one child, and some families may also include grandparents. The relatively low number of cases in these home outbreaks might be considered an advantage of compulsory home quarantine because transmission was limited to the small number of family members. Similar stay-at-home policies have now been implemented elsewhere during the pandemic.

The rising trend shown before the peak period in Figure 2 was probably due to the introduction of imported cases due to the Spring Festival travel season (Chunyun in Chinese), a period around CNY during which many people leave cities in which they work to visit their rural families. The 2020 Chunyun brought people from the epicentre Wuhan to their home cities before Wuhan’s lockdown on 23 January. Social and family gatherings continued after 23 January in most cities outside Hubei. Interventions such as contact tracing and confinement of estates, villages, and individual buildings were implemented gradually in most cities outside Hubei immediately after CNY, which explains the sharp declining curve after 28 January.

Our study does not rule out outdoor transmission of the virus. However, among our 7,324 identified cases in China with sufficient descriptions, only one outdoor outbreak involving two cases occurred in a village in Shangqiu, Henan. A 27-year-old man had a conversation outdoors with an individual who had returned from Wuhan on 25 January and had the onset of symptoms on 1 February.

The second salient feature of the 318 identified outbreaks is the relatively small number of outbreaks that involved 10 or more cases. The largest outbreak occurred in a Tianjin shopping mall and involved 21 cases, although Wu et al.^5^ reported that 25 cases were involved (Table 1). This feature contrasts with the 2003 SARS-CoV epidemic, during which 7 major super-spreading events in Hong Kong and Singapore alone were identified to involve as many as 329 cases, and super-spreading events dominated the epidemic.^6^ The occurrence of many small outbreaks (in number of cases) in the COVID-19 pandemic suggests a different transmission pattern from that of the 2003 SARS-CoV epidemic. Some virus, epidemiological, and environmental factors could have contributed to this observed difference between the 2003 SARS-CoV epidemic and the current COVID-19 pandemic.

We cannot pinpoint the exact transmission routes from these identified outbreaks. Most health authorities advised that the COVID-19 virus is transmitted mainly by close contact and via the fomite route (e.g., China NHC^7^ and CDC^8^). The China NHC also suggested that long-range aerosol transmission may occur when certain conditions are met, such as in crowded enclosures or spaces with poor ventilation. Frequent close contact occurs and high touch surfaces exist in buildings.^9–12^ We do not have data on the hygiene conditions and human density of the infection venues of the 318 outbreaks studied here. The exact location of the infection venues and the necessary parameters such as the floor area or the number of occupants were not provided in the case reports. Instead, we reviewed the current design standards of thermal and ventilation conditions, occupant density and close contact behaviour in the various indoor environments discussed here (Table S3). The required ventilation rates vary significantly among homes, offices, trains, and buses. For example, the required ventilation rate is only 3·9 L/s per person in shopping malls and 2·8 L/s per person in public buses, whereas a ventilation rate of 8 to 10 L/s is required for good indoor air quality.^26^ An international systematic review showed that a rate as high as 25 L/s per person may be needed.^13^ Many existing buildings are crowded, poorly ventilated, and unhygienic. A comprehensive review of ventilation conditions in Chinese indoor environments by Ye et al.^14^ showed that the CO_2_ concentration can reach 3,500 ppm in some buildings. The design and operation of buildings have also been under pressure to reduce energy use^15^ and increase human productivity. Balancing the need for energy efficiency, indoor environment, and health in both urban planning and building design has not been easy.^16^ The quality of indoor environments might be sacrificed by putting a greater focus on cost than on health.

This study has limitations. We only studied outbreaks in China, where very strict intervention measures were implemented. We relied fully on the case reports of the local health authorities in each city, and variation exists in the details and the quality of their original epidemiological investigations. We also made no attempt to access any of the infection venues, and the details of each of these indoor spaces remains unknown.

This study shows that the individual indoor environments in which we live and work are the most common venues in which the virus of the once-in-a-century-pandemic is transmitted among us. An individual infected in one building may infect others in the building(s) that he or she later visits. People are in constant contact as they move from one indoor space or building to another, which creates an indoor contact network through which a virus can spread.^17^ The buildings and transport cabins in various parts of the world are thus connected and facilitated the spread of the COVID-19 pandemic virus.

The association between crowding and infection has been known since Pringle.^18^ The most dramatic example might be in the cruise ship outbreak on the crowded Diamond Princess, of which the peak basic production number was predicted to be 11^19^ or 14·8^20^ before quarantine and much higher elsewhere. The world’s first statutory housing policy, the Artisans and Labourers Dwellings Act 1875^21^ was developed following 19^th^ century empirical evidence that crowding led to a high incidence of infectious disease. A recent systematic review by the World Health Organization also found an association between crowding and infection.^22^ A *Lancet* editorial in 2018 declared ‘[t]he right to a healthy home’.^23^ One WHO meeting also declared that ‘everyone has the right to breathe healthy indoor air’^24^ and that ‘the provision of healthy indoor air should not compromise global or local ecological integrity, or the rights of future generations’.^25^ We hope that in the post-pandemic future, mankind will reflect deeply on the need for a healthy indoor environment.

## Data Availability

Not available

## Acknowledgments

The authors are grateful to Shengqi Wang, Xiaoxue Cheng, Luping Ma, Ya Lei, Siying Mei, Ziying Zhou, Yiran Lu, Mengjie Duan, Yifan Li, Qionglan He and Ziang Xi, who helped collect the original data.

## Authors’ contribution

The authors declare no conflict of interest.

H. Qian, T. Miao, and Y. Li contributed equally. Y. Li and H. Qian contributed to study design, hypothesis formulation, and coordination. L. Liu, X. Zheng and D. Luo contributed to data collection and initial data analyses. H. Qian and T Miao contributed to major data analyses. Y. Li wrote the first draft of the paper, and H. Qian and T. Miao contributed to major revision. All of the other authors contributed to revision.

All of the authors approved the submitted version and have agreed to be personally accountable for their own contributions.

## Supplementary Information

**Figure S1.**
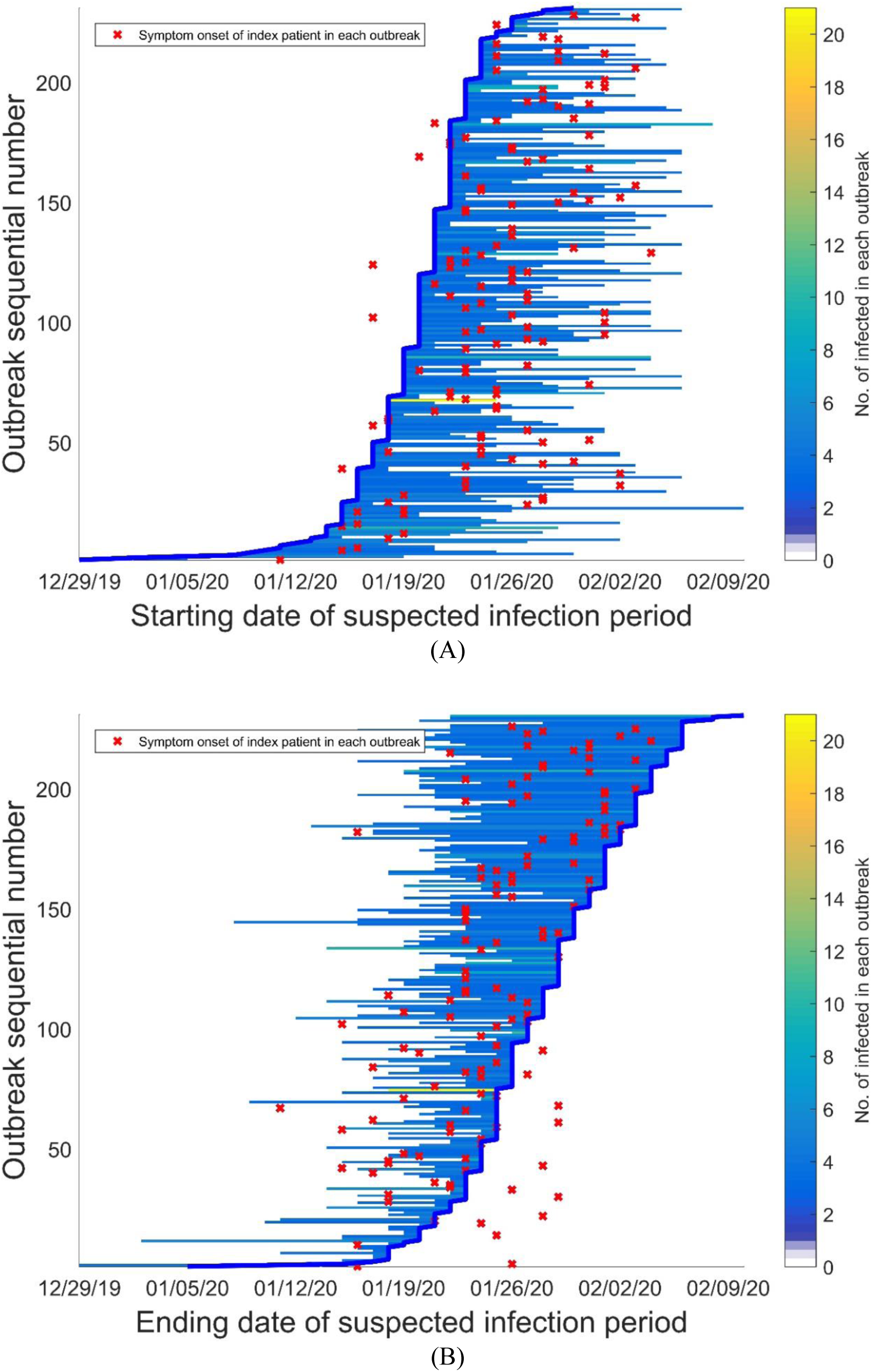
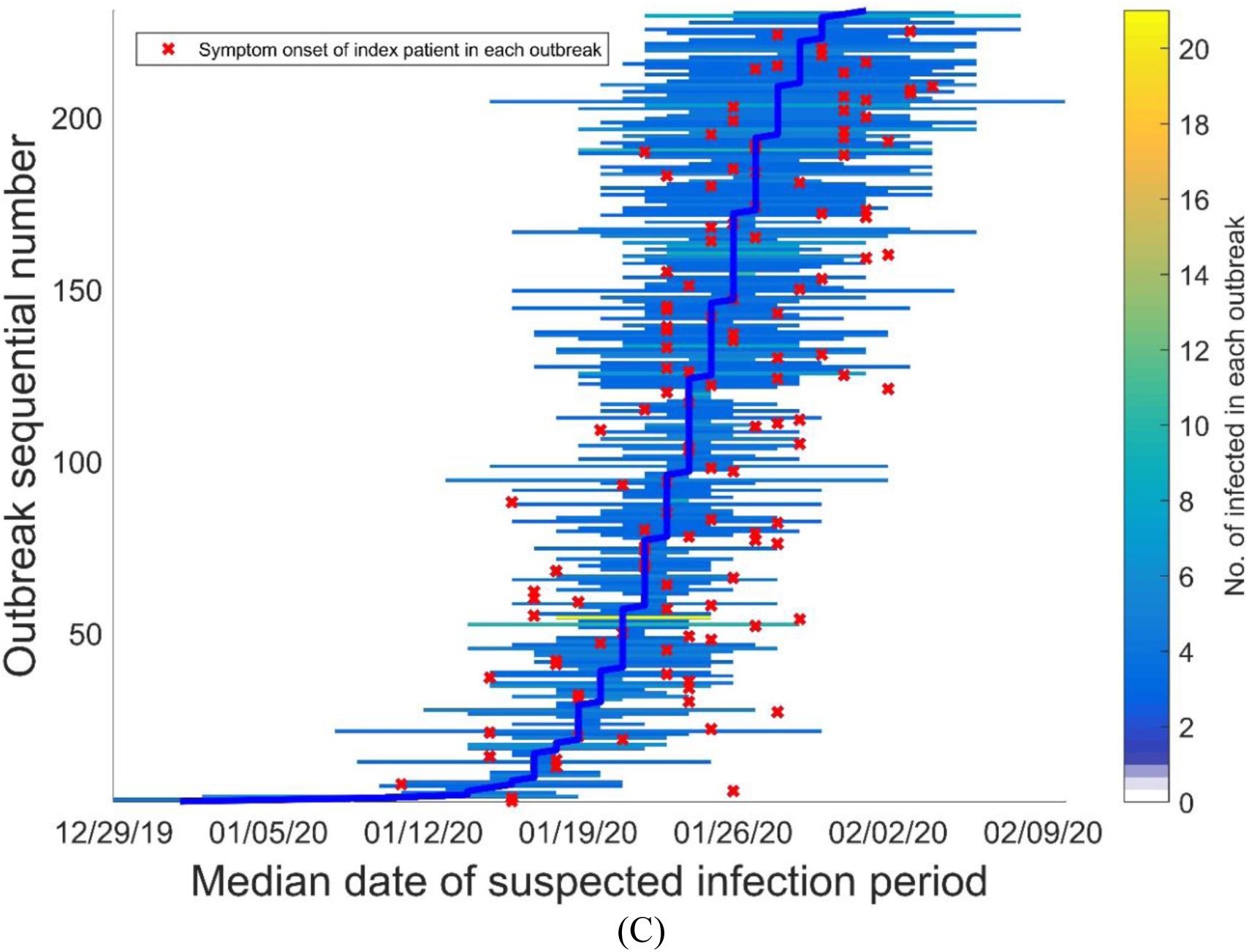
Two hundred and thirty-one outbreaks with known starting and ending dates of suspected infectious period, arranged by (A) starting date of suspected infectious period, (B) ending date, and (C) median date.

**Figure S2.**
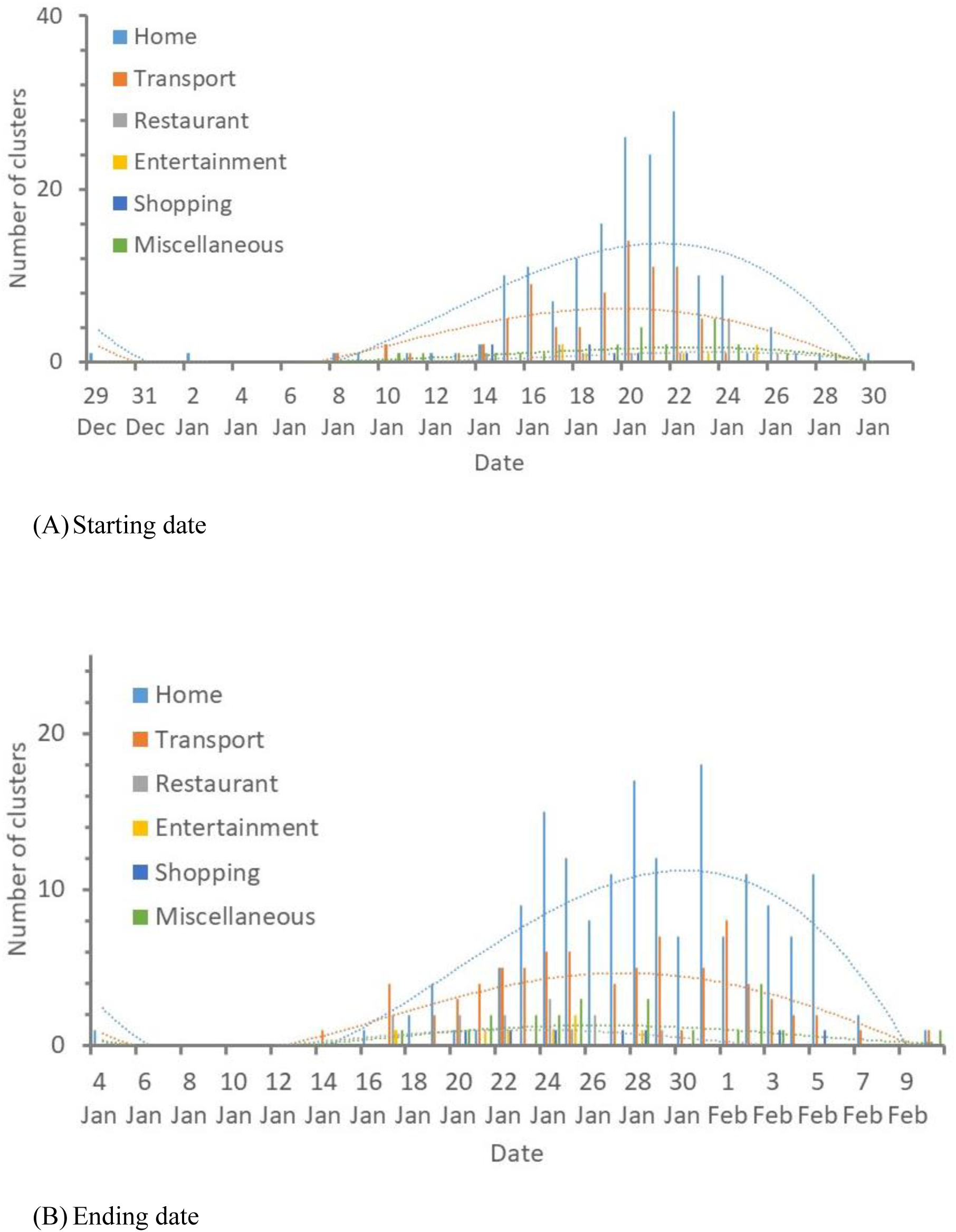
The occurrence of outbreaks in different venues on (A) starting dates and (B) ending dates of the infectious period for the 231 actual outbreaks and 300 occurred venues.

**Figure S3.**
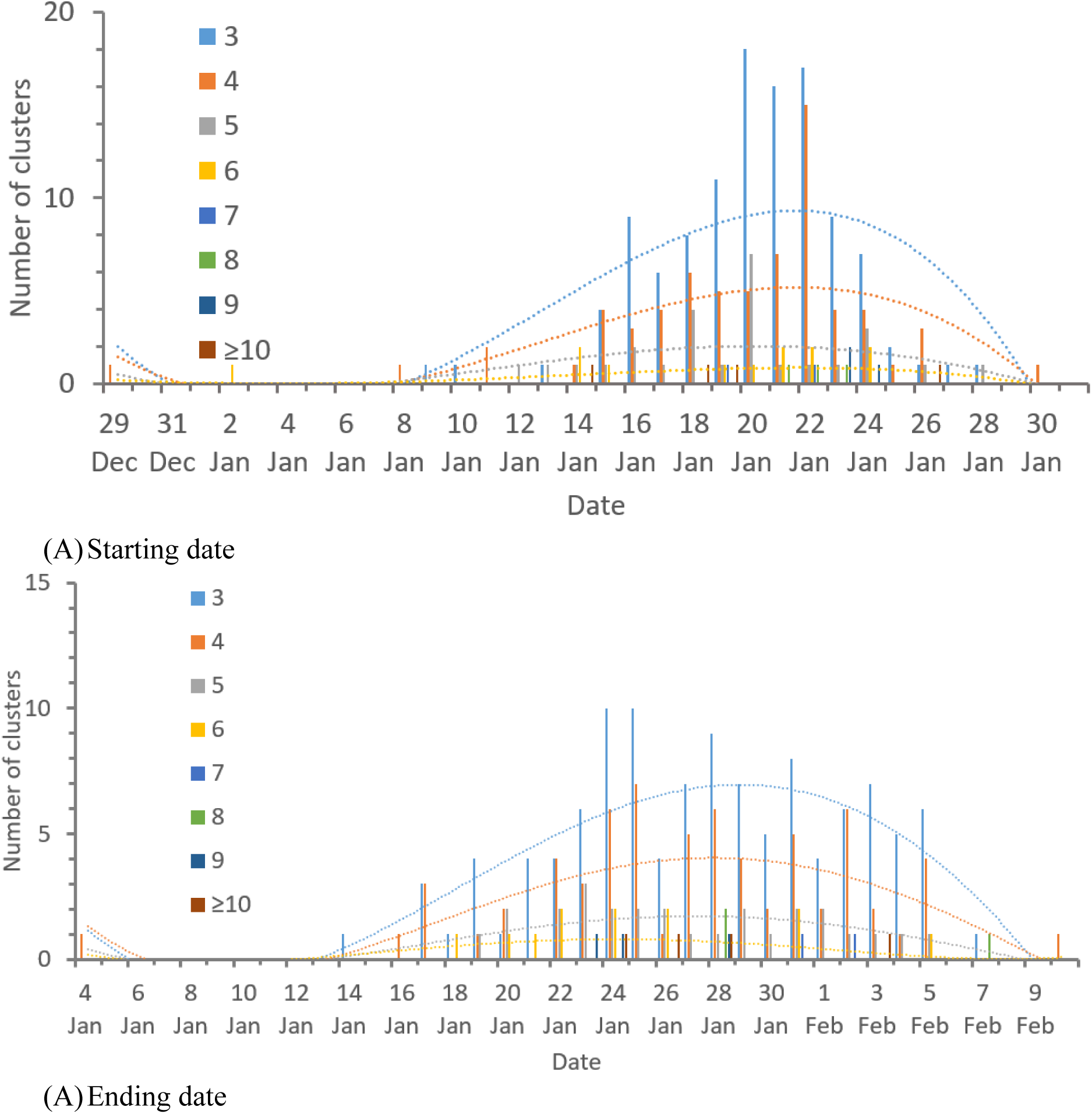
Occurrence of outbreaks with numbers of cases in each outbreak on (A) starting dates and (B) ending dates of infectious period for 231 actual outbreaks.

**Figure S4.**
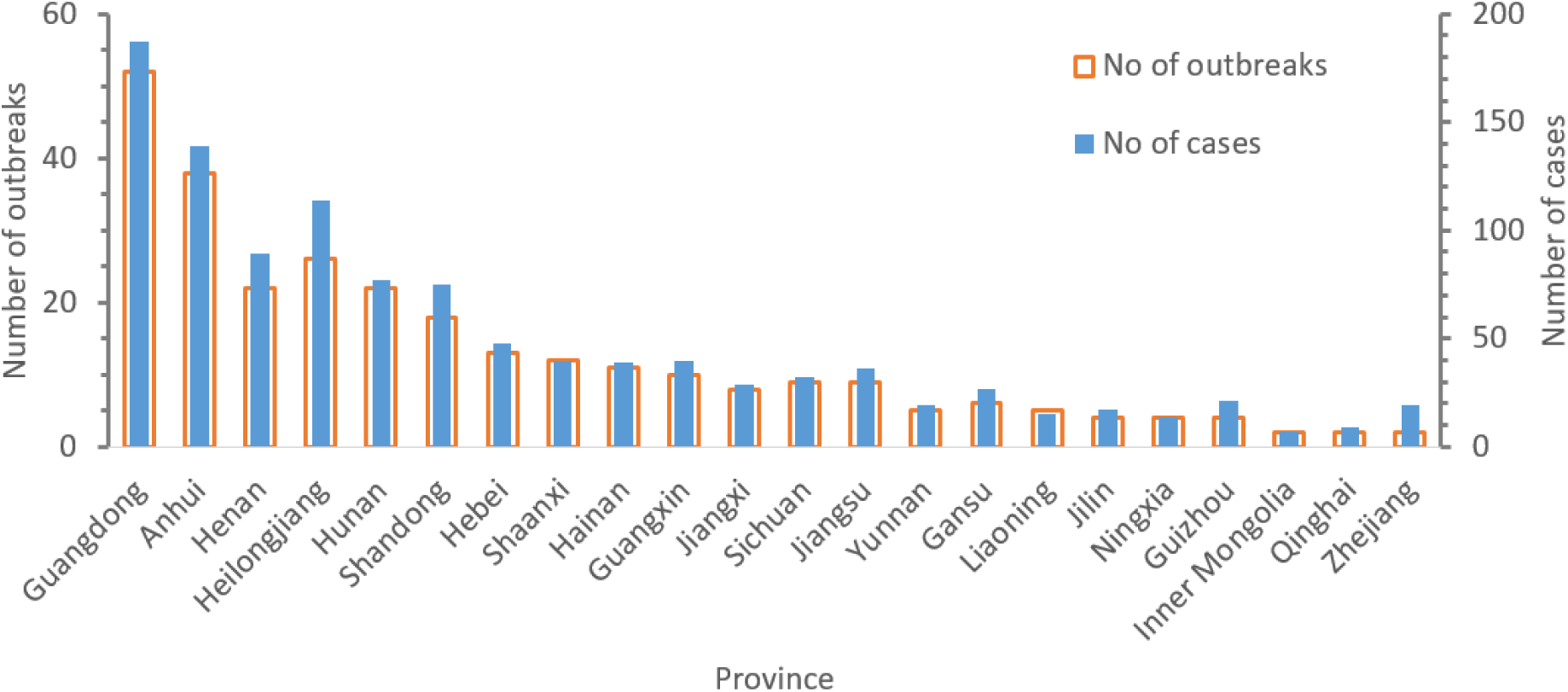
Provincial distribution of the identified outbreaks and number of cases involved in these outbreaks.

**Table S1.** Brief summary of 4 outbreaks with 10 to 21 cases.

| Indoor category (cases) | Brief description |
| --- | --- |
| Shop (10), Liaocheng, Shangdong | Between 30 January and 7 February, 10 workers from the Zhenhua Supermarket (振华量贩聊城闸口店) were confirmed to be infected. Seven of them worked in the 1F shopping area. Six other secondary infections were reported (five family members of the two cases and one close contact of the third case). Local health authorities identified 169 close contacts by 11 February, and a free check and test were offered to all customers who visited the shop from 16 to 30 Jan.<br>Additional reference<br>China Economic Weekly 中国经济周刊. Feb 19. 山东一超市负责人瞒报致 17 人确诊 169 人隔离. The person in charge of a supermarket in Shandong concealed the report leading to 17 cases confirmed and 169 people quarantined. https://finance.sina.com.cn/china/gncj/2020-02-19/doc-iimxyqvz4206652.shtml; accessed March 28, 2020. |
| Family gathering (11), Hong Kong | Nineteen members of three families, including two relatives from Guangdong, who attended a hotpot family gathering at Lento Party Room on **26 January**. Eleven were infected; the first case showed symptoms on 30 January. The venue has a hotpot area, BBQ space, mahjong table, ping pong table, pool table, and karaoke. |
| Shop (10), Wenzhou, Zhejiang | Based on our collected data, 10 cases were involved. According to news reports (Wenzhou News Network, 2020), a total of 17 cases were involved in this outbreak, including 5 administrators, 5 shopping assistants or cleaners, and 7 customers of Wenzhou Intime Department Store (温州银泰百货商场). The patient with the earliest onset of |
|  | symptoms was a female cleaner (16 January) and the last was a female customer (1 February). The first case was identified on 20 January. The mall was closed on 22 January. The local health authority traced more than 100,000 people who visited the mall or nearby for 15 minutes or more, including 1,330 workers in the mall and nearby residents. The mall was opened again on 5 March.<br>Additional references<br>Wenzhou News Network 温州新闻网. Feb 22. 经典战“疫”：关乎 10 万人的温州鹿城“银泰”保卫战. Feb 22. The classic battle ‘epidemic’: Wenzhou Lucheng ‘Yintai’: A defense battle about 100,000 people. Accessed on March 22, 2020. http://news.66wz.com/system/2020/02/22/105238333.shtml<br>WEMP. Mar 5. 银泰温州世贸店开门了！现场如何？. The Intime Wenzhou World Trade Store opens! How about the scene? https://wemp.app/posts/58ae0542-dd75-46e5-8da1-039abf00bc4d. Accessed on March 22, 2020. |
| Shop (21),<br>Tianjin | Based on our collected data, 21 cases were involved. According to Wu et al. (2020), 25 cases were involved. Six shop workers and nine customers in a shopping centre were infected, with onset dates between 20 January (a shop assistant) and 3 February (a customer). The shopping complex was built in 1981, with a 4,000 m<sup>2</sup> shopping area, mainly on the first and second floors. There were 213 workers including shop assistants. People were purchasing clothes and gifts before CNY, so the shop was very crowded with at least 10,000 visitors to the venue during the possible infectious periods. The outbreak mainly occurred on the first floor, where clothes, shoes, jewellery, and small appliances are sold. The building was closed on 26 January.<br>Additional reference<br>Wu W, Li Y, Wei Z, et al. Investigation and analysis on characteristics of a cluster of COVID-19 associated with exposure in a department store in Tianjin. Chin J Epidemiol. April 2020 vol 41 No 4 pp. 489-493. |

**Table S2.** Distribution of the identified outbreaks in 120 cities.

| No | City in Chinese | City in English | Number of outbreaks | Number of cases involved in the identified outbreaks |
| --- | --- | --- | --- | --- |
| 1 | 深圳 | Shenzhen | 24 | 84 |
| 2 | 重庆 | Chongqing | 16 | 61 |
| 3 | 亳州 | Bozhou | 9 | 35 |
| 4 | 岳阳 | Yueyang | 9 | 29 |
| 5 | 阜阳 | Fuyang | 8 | 27 |
| 6 | 蚌埠 | Bengbu | 7 | 25 |
| 7 | 天津 | Tianjing | 7 | 46 |
| 8 | 鸡西 | Jixi | 6 | 21 |
| 9 | 新乡 | Xinxiang | 6 | 20 |
| 10 | 海口 | Haikou | 6 | 23 |
| 11 | 济南 | Jinan | 6 | 26 |
| 12 | 邵阳 | Shaoyang | 6 | 24 |
| 13 | 惠州 | Huizhou | 5 | 22 |
| 14 | 郑州 | Zhengzhou | 5 | 25 |
| 15 | 西安 | Xi'an | 5 | 15 |
| 16 | 长春 | Changchun | 4 | 17 |
| 17 | 唐山 | Tangshan | 4 | 14 |
| 18 | 东莞 | Dongguan | 4 | 13 |
| 19 | 中山 | Zhongshan | 4 | 13 |
| 20 | 哈尔滨 | Harbin | 4 | 16 |
| 21 | 赣州 | Ganzhou | 4 | 12 |
| 22 | 三亚 | Sanya | 4 | 13 |
| 23 | 常州 | Changzhou | 4 | 16 |
| 24 | 烟台 | Yantai | 4 | 15 |
| 25 | 南宁 | Nanning | 3 | 13 |
| 26 | 安庆 | Anqing | 3 | 9 |
| 27 | 六安 | Liuan | 3 | 13 |
| 28 | 铜陵 | Tongling | 3 | 9 |
| 29 | 兰州 | Lanzhou | 3 | 15 |
| 30 | 香港 | Hongkong | 3 | 17 |
| 31 | 佛山 | Foshan | 3 | 11 |
| 32 | 佳木斯 | Jiamusi | 3 | 11 |
| 33 | 七台河 | Qitaihe | 3 | 9 |
| 34 | 抚州 | Fuzhou | 3 | 13 |
| 35 | 沈阳 | Shenyang | 3 | 9 |
| 36 | 枣庄 | Zaozhuang | 3 | 10 |
| 37 | 常德 | Changde | 3 | 11 |
| 38 | 昆明 | Kunming | 2 | 6 |
| 39 | 北海 | Beihai | 2 | 6 |
| 40 | 柳州 | Liuzhou | 2 | 7 |
| 41 | 西宁 | Xining | 2 | 9 |
| 42 | 马鞍山 | Maanshan | 2 | 9 |
| 43 | 石家庄 | Shijiazhuang | 2 | 8 |
| 44 | 张家口 | Zhangjiakou | 2 | 8 |
| 45 | 珠海 | Zhuhai | 2 | 7 |
| 46 | 汕头 | Shantou | 2 | 7 |
| 47 | 肇庆 | Zhaoqing | 2 | 7 |
| 48 | 吴忠 | Wuzhong | 2 | 6 |
| 49 | 大庆 | Daqing | 2 | 10 |
| 50 | 牡丹江 | Mudanjiang | 2 | 9 |
| 51 | 齐齐哈尔 | Qiqihar | 2 | 15 |
| 52 | 绥化 | Suihua | 2 | 14 |
| 53 | 信阳 | Xinyang | 2 | 9 |
| 54 | 周口 | Zhoukou | 2 | 7 |
| 55 | 洛阳 | Luoyang | 2 | 6 |
| 56 | 内江 | Neijiang | 2 | 7 |
| 57 | 德阳 | Deyang | 2 | 7 |
| 58 | 攀枝花 | Panzhuhua | 2 | 9 |
| 59 | 淮安 | Huaian | 2 | 6 |
| 60 | 徐州 | Xuzhou | 2 | 10 |
| 61 | 铜川 | Tongchuan | 2 | 9 |
| 62 | 怀化 | Huaihua | 2 | 6 |
| 63 | 湘潭 | Xiangtan | 2 | 7 |
| 64 | 遵义 | Zunyi | 2 | 8 |
| 65 | 玉溪 | Yuxi | 1 | 7 |
| 66 | 西双版纳 | Xishuangbanna | 1 | 3 |
| 67 | 大理 | Dali | 1 | 3 |
| 68 | 河池 | Hechi | 1 | 6 |
| 69 | 贵港 | Guigang | 1 | 4 |
| 70 | 钦州 | Qinzhou | 1 | 4 |
| 71 | 合肥 | Hefei | 1 | 5 |
| 72 | 淮北 | Huaipei | 1 | 4 |
| 73 | 宿州 | Suzhou | 1 | 3 |
| 74 | 沧州 | Cangzhou | 1 | 4 |
| 75 | 邢台 | Xingtai | 1 | 4 |
| 76 | 保定 | Baoding | 1 | 3 |
| 77 | 廊坊 | Langfang | 1 | 3 |
| 78 | 承德 | Chengde | 1 | 4 |
| 79 | 陇南 | Longnan | 1 | 3 |
| 80 | 甘南州 | Gannanzhou | 1 | 5 |
| 81 | 平凉 | Pingliang | 1 | 4 |
| 82 | 梅州 | Meizhou | 1 | 3 |
| 83 | 江门 | Jiangmen | 1 | 4 |
| 84 | 韶关 | Shaoguan | 1 | 5 |
| 85 | 茂名 | Maoming | 1 | 3 |
| 86 | 汕尾 | Shanwei | 1 | 3 |
| 87 | 潮州 | Chaozhou | 1 | 5 |
| 88 | 宁夏 | Ningxia | 1 | 5 |
| 89 | 银川 | Yinchuan | 1 | 3 |
| 90 | 呼和浩特 | Hohhot | 1 | 4 |
| 91 | 鄂尔多斯 | Ordos | 1 | 3 |
| 92 | 双鸭山 | Shuangyashan | 1 | 4 |
| 93 | 黑河 | Heihe | 1 | 5 |
| 94 | 驻马店 | Zhumadian | 1 | 6 |
| 95 | 商丘 | Shangqiu | 1 | 4 |
| 96 | 漯河 | Luohe | 1 | 3 |
| 97 | 开封 | Kaifeng | 1 | 5 |
| 98 | 鹤壁 | Hebi | 1 | 4 |
| 99 | 鹰潭 | Yingtian | 1 | 4 |
| 100 | 儋州 | Danzhou | 1 | 3 |
| 101 | 大连 | Dalian | 1 | 3 |
| 102 | 朝阳 | Chaoyang | 1 | 3 |
| 103 | 广安 | Guang'an | 1 | 3 |
| 104 | 泸州 | Luzhou | 1 | 3 |
| 105 | 广元 | Guangyuan | 1 | 3 |
| 106 | 杭州 | Hangzhou | 1 | 9 |
| 107 | 温州 | Wenzhou | 1 | 10 |
| 108 | 南京 | Nanjing | 1 | 4 |
| 109 | 青岛 | Qingdao | 1 | 4 |
| 110 | 淄博 | Zibo | 1 | 4 |
| 111 | 聊城 | Liaocheng | 1 | 10 |
| 112 | 滨州 | Binzhou | 1 | 3 |
| 113 | 菏泽 | Heze | 1 | 3 |
| 114 | 汉中 | Hanzhong | 1 | 3 |
| 115 | 延安 | Zibo | 1 | 4 |
| 116 | 渭南 | Weinan | 1 | 3 |
| 117 | 商洛 | Shangluo | 1 | 3 |
| 118 | 安康 | Ankang | 1 | 3 |
| 119 | 贵阳 | Guiyang | 1 | 5 |
| 120 | 黔南 | Qiannan | 1 | 8 |
| 121 | 不确定 | Undetermined | 8 | 28 |
| Total |  |  | 318 | 1245 |

**Table S3.** Characteristics of indoor environments as required by standards where COVID-19 clusters occurred.

| Indoor | Thermal conditions | Ventilation rate (L/s.person) (measured CO2 level ppm range) | Occupant density (no/m<sup>2</sup>) | Close contact behaviour (international personal distance) | References |
| --- | --- | --- | --- | --- | --- |
| Home | 18°C-24°C, 30%-60% RH* | 5.8 (or 0.7 ACH for ≤10 m<sup>2</sup>) Median 0.34 ACH (Hou et al., 2019) |  | intimate 0.32 m | GB50736-2012 Sorokowska et al. (2017) |
| Office |  | 8.5 | 0.05 | 0.67 m | ASHRAE62.1-2004 Zhang et al. (2019) |
| Shopping centre |  | 3.9 (GB) 7.8 (ASHRAE) | 0.30-0.60 (GB) 0.15 (ASHRAE) | social 1.35 m | GB50736-2012 GB50016-2014 ASHRAE62.1-2004 Sorokowska et al. (2017) |
| Supermarket |  | 3.9 (GB) 7.6 (ASHRAE) | 0.08 (ASHRAE) |  | GB50736-2012 ASHRAE62.1-2004 |
| Kiosk |  | 7.8 (ASHRAE) | 0.15 (ASHRAE) |  | ASHRAE62.1-2004 |
| Entertainment (e.g., mahjong, dance floor) |  | 10.3 (ASHRAE) (800-2500 ppm) *<sup>2</sup> | 1.0 (ASHRAE) | intimate 0.32 m | ASHRAE62.1-2004 |
| Health club (aerobics) |  | 10 (GB) 10.8 (ASHRAE) (1750-3500 ppm) *<sup>2</sup> | 0.4 (ASHRAE) | social 1.35 m | GB50736-2012 ASHRAE62.1-2004 |
| Restaurant | 16°C-22°C, ≥20% RH (winter) 24°C-28°C, ≤60% RH (summer) | 4.1 (GB) or 6.4-6.9 (JGJ) 5.1 (ASHRAE) (500-900 ppm) *<sup>2</sup> | 0.8-1.0 (JGJ) 0.7 (ASHRAE) | intimate 0.32 m +social 1.35 m | GB50736-2012 JGJ64-2017 ASHRAE62.1-2004 |
| Hotel room |  | 8.3 (GB) 5.5 (ASHRAE) (400-1000 ppm) *<sup>2</sup> | 0.1 (ASHRAE) |  | GB50736-2012 ASHRAE62.1-2004 |
| Meeting rooms |  | 2.5 (GB) 3.1 (ASHRAE) | 0.5 (ASHRAE) | social 1.35 m | GB50736-2012 Sorokowska et al. (2017) |
| Cruise ship | 22°C (winter) 27°C, 50% RH (summer) | 8 >40% of the total air |  |  | BS EN ISO7527:2004 |
| Train and high-speed rail cabins | 26°C-28°C (summer) 18°C-20°C (winter) 40%-80% RH | 1500 ppm |  |  | TB/T1932-2014 |
| Passenger planes |  | 4.7 (at 2438 m) (FAR) 3.5 in air (ASHRAE) 9.4 on ground (ASHRAE) |  |  | FAR14CFR25.831 ANSI/ASHRAR16 1-2007 |
| Public bus | <30°C when outdoor 38°C<br>>5°C (7 for new bus)<br>(Summer);<br>≥12°C<br>(Winter) | 2.8 | 8 |  | CJ/T134-2001<br>GB7258-2012 |
| Metro and metro station | Metro station:<br><30°C;<br>40%-70% | Metro station:<br>8.3 (ventilation system on)<br>3.5 (off or when using AC)<br>Subway train:<br>2.8 with AC<br>5.6 with mechanical ventilation | Train: 5 |  | GB50157-2013<br>DB11/ 995-2013<br>GB/T7928-2003 |

\*1) For estimation of ventilation rates in standards when air change rate (ACH) is specified, we consider a room to be 3 m high.
\*2) Ye W, Wang H, Zhang X. Preliminary discussion on ventilation rates for public buildings in China. Proceedings of COBEE 2018. 2018 Feb 5-9, Austin.
In general, we tried to review the Chinese Standards/studies of Chinese buildings and cited international standards or studies elsewhere if needed.
References (all standards can be searched using the standard reference number as in the table).
- Cheng PL, Li X. Air infiltration rates in the bedrooms of 202 residences and estimated parametric infiltration rate distribution in Guangzhou, China. *Energ Buildings* 2018; **164**: 219–25. - Hou J, Sun Y, Chen Q, et al. Air change rates in urban Chinese bedrooms. *Indoor Air* 2019; **29**(5): 828–39. - Zhang J, Klepac P, Read JM, et al. Patterns of human social contact and contact with animals in Shanghai, China. *Sci Rep* 2019; **9**(1): 1–11. - Read JM, Lessler J, Riley S, et al. Social mixing patterns in rural and urban areas of southern China. *Proc Roy Soc B Biol Sci* 2014; **281**: 20140268. - Sorokowska A, Sorokowski P, Hilpert P, et al. Preferred interpersonal distances: a global comparison. *J Cross-Cult Psychol* 2017; **48**:577–92.
Cheng and Li (2018) found that only 16% of bedrooms satisfied the ventilation requirement. Hou et al. (2019) stated ‘The result is that in approximately 54% of Chinese bedrooms, regardless of climate region or season, the only outdoor air entering bedrooms is by infiltration, and infiltration rates are low (median: 0.34 h<sup>-1</sup>)’.

